# The Neural Correlates of Alcohol Approach Bias – New Insights from a Whole-Brain Network Analysis Perspective

**DOI:** 10.1101/2024.09.26.24314399

**Authors:** Angela M. Muller, Victoria Manning, Christy Y.F. Wong, David L. Pennington

## Abstract

Alcohol approach bias, a tendency to approach rather than to avoid alcohol and alcohol-related cues regardless of associated negative consequences, is an emerging key characteristic of alcohol use disorder (AUD). Reaction times from the Approach-Avoidance Task (AAT) can be used to quantify alcohol approach bias. However, only a handful of studies have investigated the neural correlates of implicit alcohol approach behavior. Graph Theory Analysis (GTA) metrics, specifically, weighted global efficiency (wGE), community detection, and inter-community information integration were used to analyze functional magnetic resonance imaging (fMRI) data of an in-scanner version of the AAT from 31 heavy drinking Veterans with AUD (HDV) engaged in out-patient treatment and 19 healthy Veterans as controls (HC). We found a functional imprint of alcohol approach bias in HDVs. HDVs showed significantly higher wGE values for approaching than for avoiding alcohol, indicating that their brain was more efficiently organized or functionally set to approach alcohol in the presence to alcohol-related external cues. In contrast, Brains of HCs did not show such a processing advantage for either the approach or avoid condition. Further post-hoc analyses revealed that HDVs and HCs differed in how they implemented top-down control when approaching/avoiding alcohol and in how the fronto-parietal control network interacted with subsystems of the default mode network. These findings contribute to understanding the complex neural underpinnings of alcohol approach bias and lay the foundation for developing more potent and targeted interventions to modify these neural patterns in AUD patients.

## 1. Introduction

One characteristic of alcohol use disorder (AUD) is an automatic behavioral tendency or bias to approach rather than to avoid alcohol despite knowledge of negative consequences and/or an explicit intention to stop alcohol use (Wiers et al., 2009). Reaction times from the Approach-Avoidance Task (AAT) can be used to quantify alcohol approach bias in AUD (Wiers et al., 2009). The AAT requires participants to use a joystick to pull (i.e., approach) or push (i.e., avoid) pictures with cues of alcohol and non-alcoholic beverages in response to a non-relevant feature of the stimuli such as the format (landscape vs portrait or left vs right tilt). The two types of beverages (alcoholic vs non-alcoholic beverages) and conditions (pull vs push) are equally distributed across all trials of the test. To create the illusion that the participant is approaching/avoiding the stimulus, the pulling/pushing movements are combined with corresponding zooming effects of the stimuli (Wiers et al., 2009). Since alcohol approach bias is an automatic action tendency, it largely eludes voluntary executive-control. To help AUD patients overcome this implicit action tendency, the approach bias modification (ApBM) training, a computerized training intervention, has been developed by adapting the original AAT (Wiers et al, 2010, Wiers et al, 2011). Instead of having to pull/push the pictures with the alcohol stimuli at an equal 50% rate, participants push away alcohol-related pictures in 90% of all instances (Wiers et al., 2009, Wiers et al., 2010). Several successful randomized controlled trials (RCT) have proven ApBM (3 -12 training sessions) to be an efficient ad-on intervention during AUD residential treatment, with patients showing reduced rates of resumption to alcohol use from 10 - 17% one year after training (Wiers et al., 2011; Eberl et al., 2013; Rinck et al., 2018; Manning et al., 2021; Salemink et al., 2021). Consequently, Australia and Germany, two of the countries in which the RCTs were conducted, have recently recommended ApBM for AUD treatment in their national clinical guidelines (Haber et al. 2021; Kiefer et al., 2021).

Despite ApBM’s efficiency for reducing resumption of alcohol use in AUD patients, only a handful of studies have tried to gain insight into the neural correlates of approach bias. Ernst et al., (2012) were the first to investigate differences in brain activity between the four AAT conditions by using functional near infrared spectroscopy (fNIRS) in 21 AUD patients and 21 controls. AUD patients showed stronger activity in orbitofrontal cortex when approaching alcohol and when avoiding non-alcohol (i.e., soda) pictures while controls showed the opposite activation pattern. The authors interpreted this finding as an indicator that AUD patients and controls assign different subjective value for approaching and avoiding alcohol and non-alcohol (Ernst et al., 2012). Controls and AUD patients both showed higher top-down control activity of the dorsolateral prefrontal cortex during the avoidance than during the approach condition. Two years later, Wiers and colleagues (2014) acquired functional magnetic resonance imaging (fMRI) data from 20 AUD patients and 17 controls while performing an MRI adapted version of the AAT. Using *a priori* defined regions of interest (ROI), they found larger BOLD signal responses in nucleus accumbens and medial prefrontal cortex in AUD patients and activation in amygdala correlated positively with alcohol craving scores during the approach contrast. All three brain regions with stronger activations in AUD patients are key regions in reward and motivation processing. The same group (Wiers et al., 2015a) also examined brain activations during the AAT in 26 AUD participants at baseline and after six ApBM (n = 13) or sham (n = 13) training sessions over 3 weeks. Before training, both groups showed significant approach bias related activation in an *a priori* defined ROI, the medial prefrontal cortex. After the training, the group that received ApBM showed a significantly stronger reduction in medial prefrontal cortex activation compared to sham group. In a final study, Wiers et al., (2015b) investigated effects on cue reactivity in 17 AUD patients who received ApBM versus 15 AUD patients who received a three-week sham training. At baseline, alcohol-related cues evoked activation in amygdala bilaterally and right nucleus accumbens. After the training, the ApBM group showed greater reductions in amygdala bilaterally than sham, and BOLD signal decreases in right amygdala activity correlated with decreases in craving in the ApBM group only.

Although all four studies provided valuable insights into the neural correlates of alcohol approach bias and ApBM training-related changes in brain activation, they all only examined a priori defined ROIs. However, complex behavior and cognition emerge from synchronized activity of groups of spatially distant brain regions temporarily forming networks or communities and are not limited only to local brain region activity. For that reason, our study aimed to use advanced network analyses (Graph Theory Analysis (GTA)) to investigate how the functional network architecture of the brain changes when the brain is approaching instead of avoiding alcohol related visual cues during the AAT and how the detected functional brain network configurations differ between AUD patients and healthy controls. In particular, we hypothesized that AUD patients’ implicit approach bias has a global whole-brain network signature that is absent among controls due to the lack of an alcohol approach tendency. AUD patients’ reaction time is typically faster when approaching alcohol cues, which may also mean that their brain is functionally more efficient for approaching than for avoiding alcohol stimuli, or in other words, their neurobiology is functionally “set to approach” alcohol stimuli. Therefore, we hypothesized that AUD patients show a significant processing advantage for approaching alcohol which can be described by the GTA measure of Global Efficiency. Weighted global efficiency quantifies the ease with which brain regions communicate and combine information from spatially distant regions (Rubinov & Sporns, 2010). Brain regions that are communicating with each other by strong functional connections (highly synchronized brain activity over time) have a higher capacity to exchange and integrate information and, as a result, are more efficiently connected than brain regions that are only weakly connected. This study additionally aimed to better understand the exact nature of this hypothesized processing advantage using post-hoc tests investigating the network/community structure and the information integration flow between these networks/communities during approaching or avoiding alcohol in AUD patients and healthy controls.

## 2. Methods

### 2.1 Participants

We describe data from 31 heavy drinking Veteran patients with AUD (HDVs) and 19 healthy control Veterans without AUD (HC) acquired during task fMRI and structural MRI. They all were part of a larger sample of Veterans participating in a study conducted at the San Francisco Veterans Affairs Medical Center (SFVAMC) to investigate the neural correlates underlying alcohol approach bias and the effects of ApMB training on the brain in this population. The study was carried out in accordance with The Code of Ethics of the World Medical Association (Declaration of Helsinki). All participants provided written informed consent prior to study and underwent procedures approved by the University of California, San Francisco and the San Francisco Veterans Affairs Health Care System (SFVAHCS). All study procedures took place at SFVAHCS in San Francisco, CA. HDVs and HCs were recruited from the San Francisco Veterans Affairs Health Care System; HDVs were engaged in an outpatient AUD treatment at the time of recruitment. Basic demographics of both groups are given in Table 1.

**Table 1.** Demographics.

|  | **HDV** | **HC** | **Statistics** |
| --- | --- | --- | --- |
| **n** | 31 | 19 |  |
| **Age (Years)** | 50.9 | 50.3 | p = .85 |
| **Education (Years)** | 13.7 | 15.9 | p < .001 |
| **Sex (Male/Female)** | 25/6 | 14/6 |  |
| **Race (n/in percent)** |  |  |  |
| White | 18/58 | 6/32 |  |
| Black/African-American | 4/13 | 3/16 |  |
| Asian | 2/7 | 6/32 |  |
| American Indian/Alaska Native | 2/7 |  |  |
| Native Hawaiian or Other Pacific Islander | 1/1 |  |  |
| Mixed Race | 4/13 | 4/21 |  |
| **Ethnicity** |  |  |  |
| Hispanic or Latino | 4/13 | 1/5 |  |
| Non-Hispanic or Non Latino | 26/84 | 17/90 |  |
| Unknown | 1/1 | 1/5 |  |
| **AUD Age of Onset (Years)** | 25.6 | – | – |
| **Drinking past 90 days** |  |  |  |
| Average of drinks/week | 31.6 | 1.3 | p < .001 |
| Average of heavy drinking day/week | 2.2 | 0 | p < .001 |
| Average of drinking day/week | 3.4 | 0.7 | p < .001 |
| **AUDIT** | 18.1 | 1.9 | p < .001 |
| **Craving** |  |  |  |
| **OCDS total** | 13.1 | 1.2 | p < .001 |
| **OCDS obsessive** | 4.9 | 0 | p < .001 |
| **OCDS compulsive** | 8.1 | 1.2 | p < .001 |

HDV = Heavy Drinking Veterans, HC = Healthy Controls, OCDS = Obsessive-Compulsive-Drinking Scale

HDVs were male and female U.S. Veterans between 18 – 65 years old, with moderate to severe AUD as defined by the Diagnostic and Statistical Manual of Mental Disorders (5^th^ ed.; DSM-5; American Psychiatric Association, 2013). In addition to meeting AUD diagnostic criteria, all HDVs reported “heavy” drinking based on National Institute on Alcohol Abuse and Alcoholism (NIAAA) criteria (i.e., at least 15 standard drinks for men and 8 drinks per week for women for at least one week in the 90 days prior to consent; Willenbring et al., 2009). All HDV participants expressed a desire to reduce or stop alcohol use and were receiving treatment for AUD at the SFVAHCS. HCs could not have exceeded heavy drinking levels in the preceding 5 years, or have a diagnosis or treatment history of AUD. Exclusion criteria for HDVs and HCs were presence of other clinically significant psychiatric disorders or unstable medical conditions that would create excessive risks; current or history of intrinsic cerebral tumors, demyelinating or neurodegenerative diseases, cerebrovascular diseases, severe or penetrating traumatic brain injury, documented learning disabilities; pregnancy or attempting to conceive; concurrent participation in another AUD study or clinical trial; and MRI contraindications. Additionally, to be included in the study presented here, participants had to have high quality structural MRI data with at least 80% of the time series (503 of in total 628) after removal of motion and physiological noise from the task fMRI data.

Alcohol use inclusion criteria were assessed using the Time Line Follow Back (TLFB; Sobell et al., 1985; Sobell & Sobell, 1992) interview, which yields total number of standard alcohol drinks per week. Alcohol craving-related obsessive thoughts and compulsions were also measured using the Obsessive Compulsive Drinking Scale (OCDS; Anton et al., 1995). The OCDS is a 14-item self-report scale that can range from 0-56, an obsessive thoughts subscale that can range from 0-24, and a compulsive drinking subscale that can range from 0-32, with higher scores indicating greater level of obsessive-compulsive drinking.

### 2.2 MRI-data

MRI data were acquired on a 3 Tesla-Magnetom Skyra Syngo MRI using a 32-channel receive head coil. The following sequences were used: (1) Structural T1 weighted images: A MPRAGE (Magnetization Prepared Rapid Acquisition Gradient Echo) sequence with repetition time (TR) 2400 ms, echo time (TE) 2.24 ms, Flip angle 8 deg, field of view (FoV) 256 with x 240 x 167 mm, isotropic voxel size 0.8 x 0.8 x 0.8 mm, slice thickness 0.8 mm, 320 slices per volume. (2) Structural T2 weighted images: A SPACE (Sampling Perfection with Application optimized Contrast using different flip angle Evolution) sequence with TR 3280 ms, TE 565 ms, Flip angle 120 deg, FoV 256 x 240 x 167 mm, isotropic voxel size 0.8 x 0.8 x 0.8 mm, slice thickness 0.8mm, 320 slices per volume (3) Task fMRI images: A whole brain echo planar imaging (EPI) sequence with TR 1000 ms, TE 36 ms, Flip angle 52 deg, multi band acceleration factor 7, FoV 224 x 224 x 140 mm, isotropic voxel size 2.0 x 2.0 x 2.0mm, 70 slices per volume, slice thickness 2mm, two sessions with 314 volumes each. (4) Fieldmaps: A sequence with TR 450 ms, TE 4.92 ms, Flip angle 40 deg, reconstruction Magn./Phase, FoV 212 x 212 x 141, voxel size 2.0 x 2.0 x 3.6mm, 42 slices.

### 2.3 fMRI version of Alcohol Approach/Avoidance Task (AAT)

NeuroBehavioral Systems Presentation software was used for visual presentation of the stimuli and for collecting patient responses. 40 alcohol and 40 non-alcohol beverage images presented against a black background were used as cues. Participants pulled/pushed an MRI-compatible Fiber Optic Joystick in response to the tilt of a cue. A right-tilt of the cue image signaled the participant to push (avoid), a left-tilt to pull (approach) the cues. Pulling the joystick resulted in a zooming effect causing the picture size to increase. Pushing the joystick resulted in an avoidance effect causing the picture size to decrease. The participants performed 12 practice trials, then 160 test trials were presented in an event-related design over 2 runs. Each run had 40 pull and 40 push trials, half of which were for alcohol and the other for non-alcohol images taken from the American Alcohol Photo Stimuli (AAPS) and the Amsterdam Beverage Picture Set (ABPS) supplemented with images of alcohol and non-alcohol beverages commonly reported to be consumed in our previous pilot study. Each of 40 alcohol and 40 non-alcohol images were presented twice during the experiment, once for the push and once for the pull condition. Participants had 2-seconds time window for completing their response to each cue. After completing a successful push/pull response, the image disappeared, followed by an intertrial interval up to 10 seconds, with a random hyperbolic distribution to optimize efficiency. Intertrial intervals accounted for varied response times preserving a fixed duration for each run. When a participant made an error (e.g., pull when cued to push) or responded too slowly (>2 seconds), a red cross appeared on the screen for 500 ms prior to being exposed to the next stimuli. Total task time was about 10 minutes.

### 2.4 Preprocessing of MRI data

SPM12 (running on MATLAB (Release 2019b, The MathWorks, Inc., Natick, Massachusetts, United States) and ANTs (Advanced Normalization Tools; Avants et al., 2011) were used for preprocessing the MRI data. (1) The “Realign & Estimate” procedure from SPM12 was used to compute the motion parameters for the EPI data. Only the resulting mean EPI image and the motion parameters were kept as input for the outlier detection and denoising procedure described in section 2.5 (2) The fMRI images were slice-time corrected and the field maps used to create voxel-displacement maps (VDM) with the SPM FieldMap toolbox. (3) The “Realign & Unwarp” procedure from SPM with the mean EPI image from step 1 as first and the VDM from step 2 was then used to create distortion and motion corrected EPI images. (4) The Coregister (Estimate & Reslice) procedure was then used for co-registering the T2w image to the T1w image as reference. (5) The distortion corrected mean EPI image from step 3 was then co-registered to the T2w image from step 4 with ANTs using the “antsRegistrationSyN.sh” procedure. (6) The ANTs transformation was then applied to all motion and distortion corrected EPI images using the “antsApplyTransformations” procedure. (7) The New Segmentation Algorithm from SPM12 was used to segment the T1w images into grey matter (GM), white matter (WM), and cerebrospinal fluid (CSF) tissue probability maps in native space and to create a rigidly aligned version of the 3 tissue maps. (8) Using the DARTEL procedure (Ashburner, 2007) from SPM12, the rigidly aligned tissue maps were used to create a study specific template and subject-specific warped images used by the normalization step. (9) Normalization of the structural and functional images to the MNI space using the “DARTEL-Normalise to MNI Space” procedure from SPM12. During that step, all images were resampled to a voxel resolution of 2 x 2 x 2 mm.

### 2.5 Creation of connectivity matrices

CONN (release 22a, Whitfield-Gabrieli & Nieto-Castanon, 2012) was used to create subject-specific connectivity matrices for each of the 4 conditions (approach alcohol, avoid alcohol, approach non-alcohol, avoid non-alcohol) of the AAT task. Except for the follow-up analyses reported in section 3.5, non-alcohol data are not included in the report. The Artifact Detection Toolbox as implemented in CONN was used to identify motion corrupted outliers (global-signal z values threshold 5, subject-motion threshold 0.9 mm) and volumes identified as outliers were censored by dummy-coding them as nuisance regressors for the denoising step. Denoising: The data were denoised using the standard pipeline implemented in CONN with regression of potential confounding effects characterized by WM timeseries (5 CompCor noise components), CSF timeseries (5 CompCor noise components), the motion and outliers linear and quadratic regressors and their first order derivatives (364 components), session and task effects and their first order derivatives (8 factors), and linear trends (2 factors) within each functional run followed by bandpass frequency filtering of the BOLD timeseries between 0.008 – 0.09 Hz. CompCor (Behzadi et al., 2007) noise components within WM and CSF were estimated by computing the average BOLD signal as well as the largest principal components orthogonal to the BOLD average within each subject’s eroded segmentation masks. The Atlas of Intrinsic Connectivity of Homotopic Areas (AICHA, Joliot et al., 2015) was used to partition the brain into 384 regions of interest (ROI) and to compute ROI-to-ROI connectivity matrices. Functional connectivity strength was represented by Fisher-transformed bivariate correlation coefficients from a weighted general-linear model, defined separately for each pair of seed and target areas, modeling the association between their BOLD signal timeseries. Individual scans were weighted by a boxcar signal characterizing each experimental condition with an SPM canonical hemodynamic response function and rectified.

### 2.5 Network analyses

The unthresholded connectivity matrices were used for the following graph theory analyses (GTA) because a) thresholding the correlations, i.e. edges or links in GTA terminology can induce spurious group differences in clinical populations (van den Heuvel et al., 2017; Hallquist & Hillary, 2018), b) weak correlations can be physiologically important (Santarnecchi et al., 2014), and c) the most dynamic connections have correlations coefficients near zero (Zalesky et al., 2014).

MATLAB scripts from the Brain Connectivity Toolbox (BCT; Rubinov & Sporns, 2010) were used to compute the GTA measurements described below.

<u>Weighted global efficiency</u> (wGE) quantifies an individual network’s ease for communication between the different regions of the brain and information integration across the network (Rubinov & Sporns, 2010). It is computed as the inverse of the average shortest path length of the network, a path being a sequence of nodes (brain regions, ROIs) and edges that connect two brain regions with each other. In weighted networks, the weighted path length is computed as the sum of individual edge lengths, which are inversely related to edge weights because large weights (strong correlations) are typical between brain regions in near proximity to each other.

<u>Community structure</u> describes the brain’s hierarchically organized architecture (Doucet et al., 2011) with its multiple levels of smaller subnetworks or communities that are nested within each other. GTA defines a community structure as a subdivision of a network into nonoverlapping groups of brain regions which maximizes the number of within-group edges and minimizes the number of between-group edges (Rubinov & Sporns, 2010).

<u>Participation coefficient</u> (PC) measures to which degree a brain region is involved in the global inter-community information integration. A brain region with a PC near 1 has a high number of connections to brain regions from other communities. When using connectivity matrices with positive and negative correlations, it is additionally possible to compute a second PC value for the negative edges which quantifies a brain region’s degree of segregation from the inter-community information exchange. A brain region with such a “negative PC” value near 1 is almost isolated from the inter-community information integration (Rubinov & Sporns, 2011). When using unthresholded weighted matrices for computing PC, brain regions always load on both kinds of PC values, i.e. they have a positive PC as well as a negative PC. To quantify a brain region’s role within the inter-community information integration with a single value, the information from the region’s “positive PC” and “negative PC” can be combined by subtracting the “negative PC” from the “positive PC”. A positive subtraction value of the thusly combined PC (combPC) indicates that the brain region’s role for the information exchange across communities is more important than its role for separating the communities from each other, whereas a negative subtraction value indicates that the brain region is mainly connected with the brain regions within its own community and not involved in inter-community information integration.

In a first step, the whole brain wGE values for each participant and each condition was computed. To determine the brain’s community structure characteristic for the approach and avoid alcohol conditions, we ran 10,000 iterations of the “modularity_louvain_und_sign” algorithm from the BCT toolbox (Rubinov & Sporns, 2010) on each participant’s individual connectivity matrix while simultaneously computing a participant specific agreement matrix, which coded how frequently two brain regions were allocated to the same community over the 10,000 iterations. Next, we computed 2 condition-specific group average agreement matrices for HDVs and HCs separately. These group-average agreement matrices were then used as input to compute group-specific and condition-specific consensus partitions using the “consensus_und” algrorithm in combination with the “community_louvain” algorithm from the BCT toolbox (Rubinov & Sporns, 2010) with 10,000 iterations. A consensus partition finds a community structure that represents the community partition that was most commonly shared by all participants and across all iterations. This procedure resulted in 4 group- and condition-specific community affiliation vectors describing how the 384 brain regions grouped when the study participants were approaching/avoiding alcohol related cues. Next, the information from these affiliation vectors was used to compute subject-specific combPC values for each of the 384 brain regions as well as the global whole brain PC value (= average value of all 384 brain regions) and the average community combPC value (= average of all ROIs allocated to the same community) for each of the detected communities.

After whole brain wGE, brain community structure, and combPC values had been established for the top hierarchy level of the brain’s nested network architecture, the same GTA measurements were then computed for the next lower hierarchy level. To do so, subject- and condition-specific sub-matrices consisting of the brain regions belonging to the same community only, were built for each community that had been detected on the top hierarchy level. Then, all the procedures described above were repeated on these new sub-matrices to better understand how efficiently and in which groupings the information exchange was organized between the brain regions within the same community. This resulted in a community-specific affiliation vector identifying the number of underlying subcommunities and the brain regions from which each of these subcommunity was composed, in addition to combPC values describing the information integration across the subcommunities within the same community and a community-specific wGE value for each of the top-hierarchy communities (see flow diagram on Figure 1). Finally, to understand to which degree the subcommunities were participating in the information integration between the top-level communities, the average combPC value for each subcommunity was computed using the nodal combPC values of the 384 brain regions from the top-level community structure.

**Figure 1.**
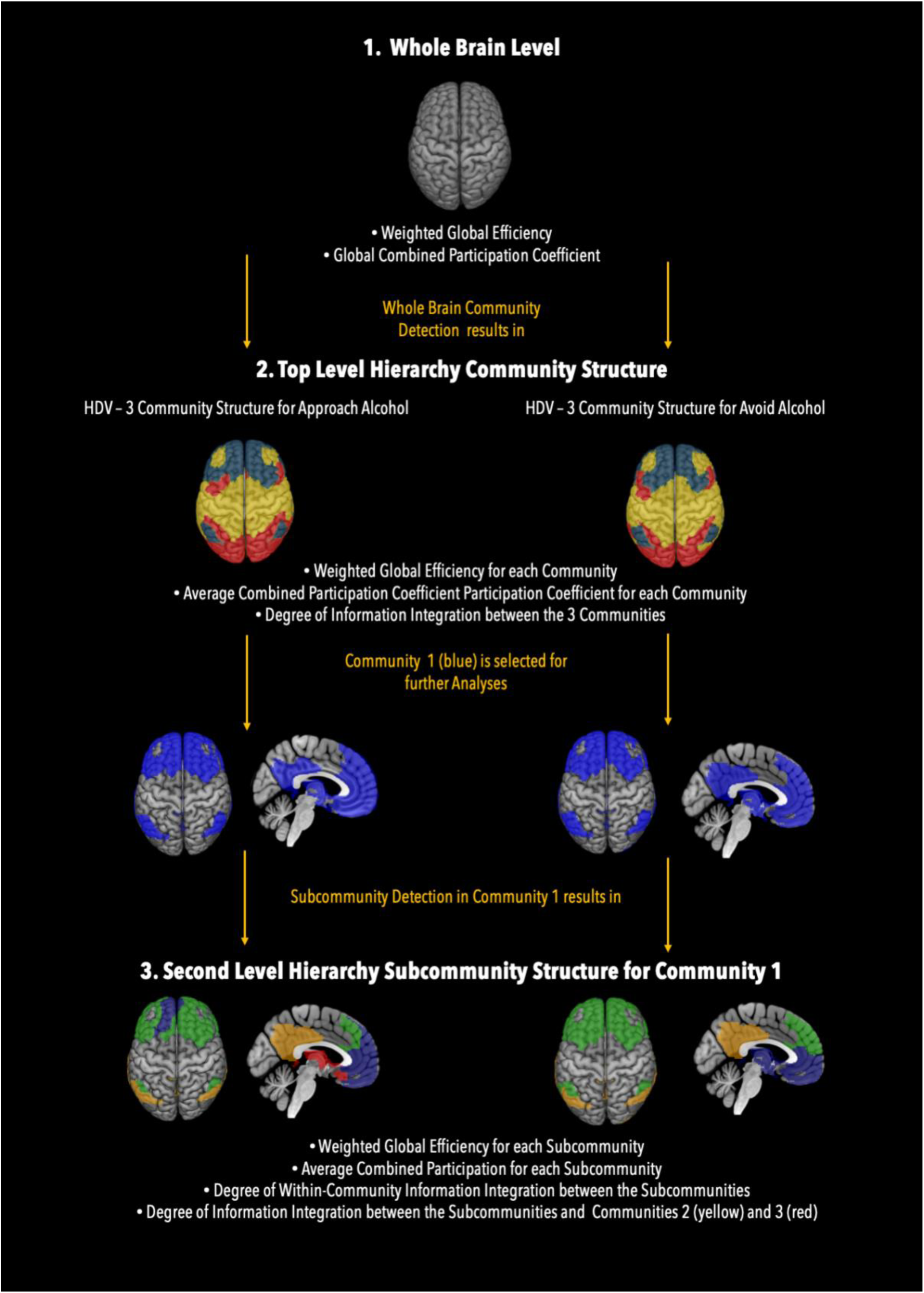
Hierarchy Levels of Analyses.

### 2.7 Statistics

The statistical software Jamovi (https://www.jamovi.org) was used to compute all the statistics. Shapiro Wilks tests were used to assess whether the used GTA measurements met the assumptions for parametric statistical tests. Weighted GE measurements were normally distributed, CombPC measurement were generally not. Consequently, Student’s t tests and non-parametric Mann-Whitney U tests were used to assess significant group differences in wGE and CombPC, respectively; paired Student T tests and signed Wilcoxon Rank Sum W tests were used to assess within-group differences between conditions; Spearman’s rho was used to test for significant associations between GTA measurements and behavior. As the primary aim of the study was to confirm our a priori hypothesis that HDVs’ functional network organization had a processing advantage for approaching alcohol, all follow-up analyses after that were treated as post-hoc tests only performed to understand which brain network characteristics were underlying the global effect. Consequently, all follow-up analyses were no longer corrected for multiple comparisons.

## 3. Results

### 3.1 Basic Demographics

Basic Demographics for the two groups are summarized in Table 1.

### 3.2 Whole brain – weighted global efficiency

HDVs’ whole brain wGE value for Approach Alcohol was significantly higher (t(30) 2.97 p = .024 (Bonferroni corrected for 4 simultaneous comparisons), Cohen’s d = 0.53) than for Avoid Alcohol indicating that the functional brain network architecture of the HDVs was significantly more efficient for approaching than avoiding alcohol or “functionally set” to approach alcohol. Across both groups and conditions, HDVs had the highest whole brain wGE value of all four computed values with 0.318 (SD 0.029) for Approach Alcohol as well as the lowest value with 0.304 (SD 0.024) for Avoid Alcohol. The HCs’ whole brain wGE values were in the middle range of the two HDVs’ values and almost identical with 0.311 (SD 0.030) for Approach Alcohol and 0.312 (SD 0.028) for Avoid Alcohol. However, whole brain wGE values between groups were not significantly different. To assess the possibility that HDV’s tendency to approach alcohol was not distinct for alcohol but rather reflected an unspecific approach impulse regardless of the object, we also computed wGE values for approaching and avoiding non-alcohol beverages. HDVs wGE value for approaching alcohol was not just the highest of all their 4 wGE values, but also significantly higher (t(30) 3.24, p = .006 uncorr, Cohen’s d = .58) than the wGE value for approach non-alcoholic beverages (0.304 (SD 0.028)). The only other significant difference (t(30), 2.12, p = .043 uncorr, Cohen’s d = .38) in wGE for the HDVs was for Avoid Alcohol vs Avoid Non-Alcohol, where the wGE value for avoiding non-alcoholic beverages (0.310 (SD 0.026) was significantly higher than for avoiding alcohol. HCs did not show significant differences in wGE for the two sorts of beverages with the exception of significantly (t(18) 2.21, p = 0.041 uncorr, Cohen’s d = .51) higher wGE values when avoiding non-alcoholic beverages (0.322 (SD 0.035) compared to avoiding alcoholic beverages.

### 3.3 Whole brain: Global combPC

HCs showed a significantly higher degree (Wilcoxon W 153, p = .018, r = .61) of inter-community information integration as measured by the global whole brain PC during Approach Alcohol when compared to Avoid Alcohol. No such between-conditions difference in global whole brain PC was found for HDVs as well as no significant between group differences in global whole brain PC (see Table 2 for details).

**Table 2.**
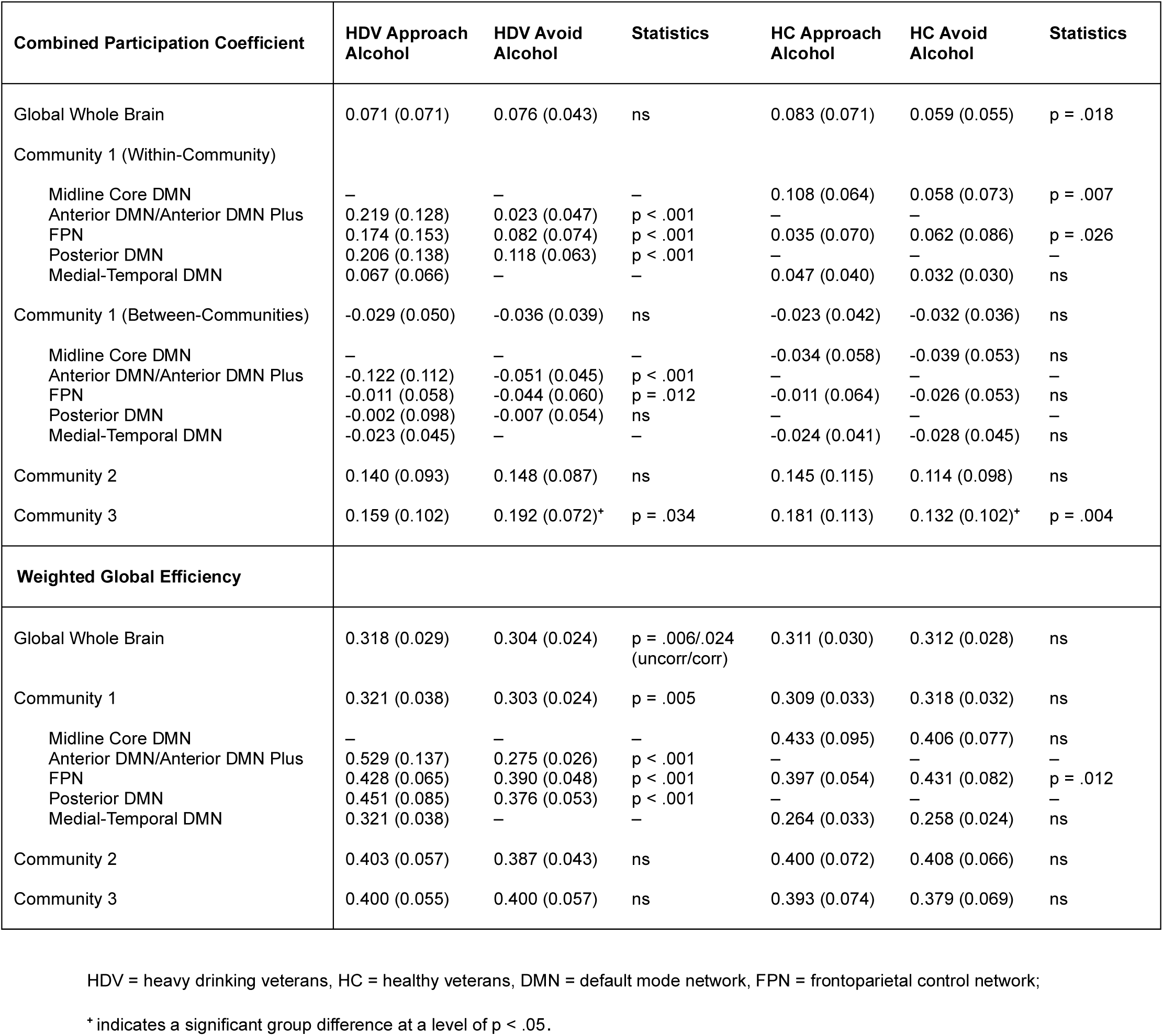
Summary of whole brain, top hierarchy and second hierarchy combined participation coefficients (combPC) and weighted global efficiency values.

### 3.4 Top Level Hierarchy: Community structure

On the top hierarchy level, both HDVs and HCs showed a bilaterally almost symmetric, three-community structure with minimal differences between conditions and groups (Figure 2). In HDVs, 32 ROIs (8%) switched community affiliation between conditions, whereas in HCs 38 ROIs (10%) changed their community affiliation when switching between conditions. The community affiliation in HDVs versus HCs differed for 50 ROIs (13%) for Approach Alcohol and for 54 ROIs (14%) for Alcohol Avoid. ROIs located in the parietal lobe (lateral and medial) and adjacent secondary visual cortex showed a tendency to switch community affiliation across groups and conditions. Community 1 (blue) comprised mainly frontal regions (bilateral, lateral and medial superior frontal gyrus, middle frontal gyrus and inferior frontal gyrus, orbital cortex, anterior cingulate cortex, ventral anterior insula) and parietal regions (bilateral angular and supramarginal gyri, inferior parietal gyrus, precuneus, posterior cingulate cortex) in combination with some lateral temporal regions (bilateral middle temporal gyrus and temporal pole), medial temporal – limbic regions (bilateral hippocampus, parahippocampal gyrus, amygdala) and subcortical brain regions, in particular nucleus caudatus and thalamus. Community 2 (yellow) primarily consisted of bilateral primary motor cortex, rolandic operculum, auditory cortex, primary somatosensory, cortex, and superior parietal lobe. Community 3 (red) encompassed the visual cortex.

**Figure 2.**
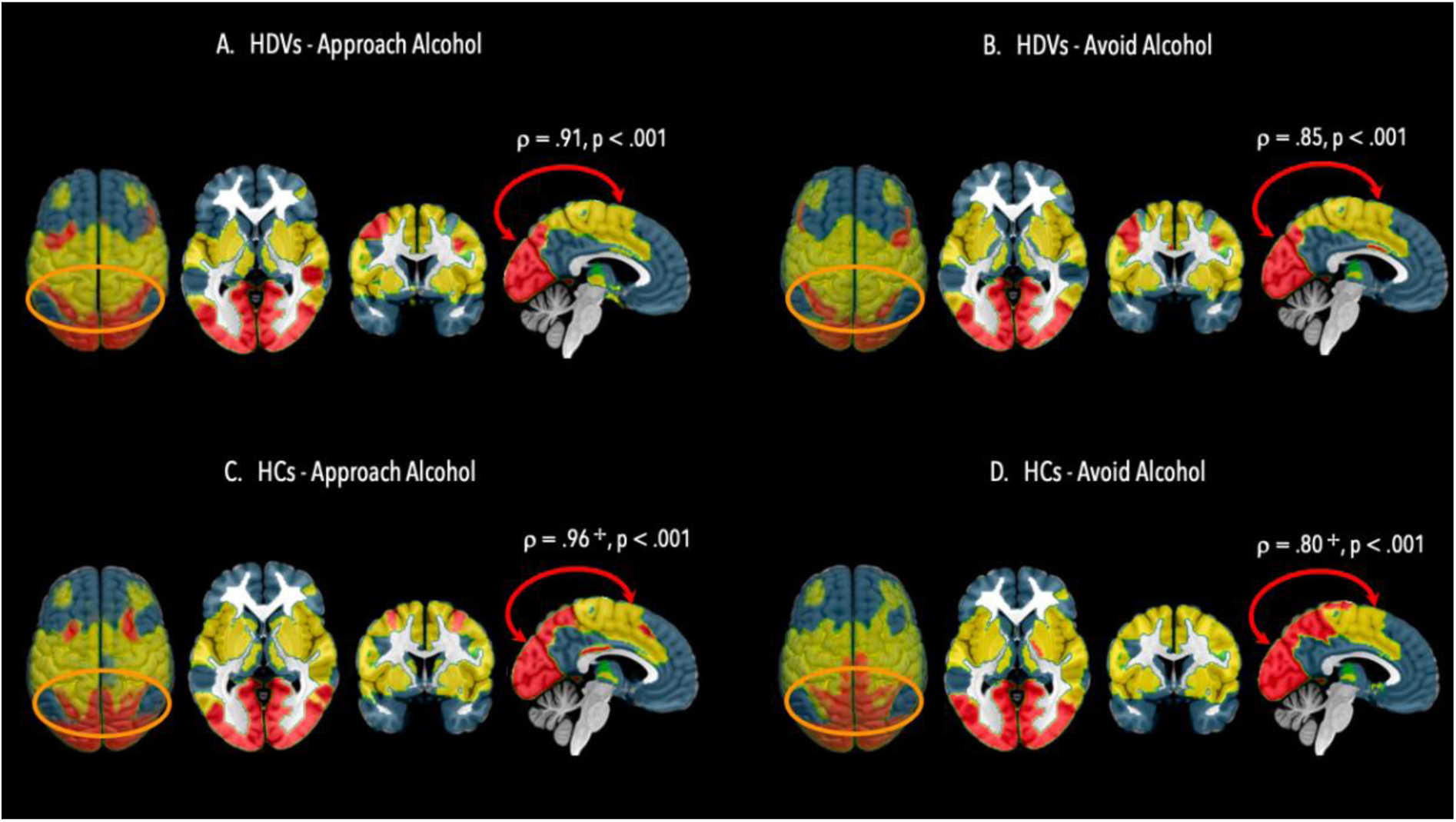
Community Structure on the Top Level Hierarchy in HDVs and HCs. Top level hierarchy community structure in heavy drinking veterans (HDV) during Approach Alcohol condition (A); Top level hierarchy community structure in HDVs during Avoid Alcohol condition (B); Top level hierarchy community structure in healthy veterans (HC) during Approach Alcohol condition (C); Top level hierarchy community structure in HC during Avoid Alcohol condition. Brain regions allocated to community 1 are in blue color and encompass brain regions belonging to the default mode network, fronto-parietal control network, brain regions allocated to community 2 are in yellow color and encompass brain regions belonging to sensorimotor network, ventral attention network and auditory-insula network, brain regions allocated to community 3 are in red color and encompass brain regions belonging dorsal attention network. Red arrows indicate significant information integration between communities, orange ovals highlight the parieto-occipital regions which showed the highest variability in community allocation across conditions and groups. + indicates a significant difference in the degree of information integration in HCs in Approach vs Avoid Alcohol conditions.

### 3.5 Top Level Hierarchy: Inter-community information integration between communities

Computing the average combPC values for the communities for each group/condition revealed that communities 2 and 3 had always positive combPC values whereas community 1 showed negative combPC values indicative of its functional segregation from the other two communities during both conditions (Table 2). Assessing the degree of inter-community information integration by computing Spearman’s rho between the average community combPC values of all possible community pairs for each condition/group separately, revealed that only communities 2 and 3 had significant inter-community information integration between each other: HDV = Approach Alcohol ρ(29) = .913, p < .001; Avoid Alcohol ρ(29) = .850, p < .001; HC = Approach Alcohol ρ(17) = .953, p < .001; Avoid Alcohol ρ(17) = .800, p < .001).

Comparing the HDVs’ global whole brain PC values and average community combPC values with the corresponding values of the HC group for Approach Alcohol revealed no significant group differences (Table 2). The findings were the same for Avoid Alcohol, i.e. no significant group differences, except for a significantly (Mann-Whitney U 160, p = .007, r = .46) higher average combPC value for community 3 in HDVs compared to HCs. Average combPC for Community 3 also showed significant differences between conditions in both groups: community 3’s average combPC value for Avoid Alcohol was significantly higher (Wilcoxon W 140, p = .034, r = -.44) as compared to Approach Alcohol in HDVs, whereas Avoid Alcohol was significantly lower (Wilcoxon W 164, p = .004, r = .73) than Approach Alcohol in HCs. The same pattern of within-condition and between-group differences that had been observed for community 3 was also present for the other two communities although not significantly. HDVs showed generally lower average combPC values for communities 2 and 3 during Approach Alcohol vs Avoid Alcohol whereas HCs showed the opposite relationship, i.e. higher average combPC values for communities 2 and 3 during Approach Alcohol vs Avoid Alcohol.

### 3.6 Top Level Hierarchy: Weighted GE of the communities

Computation of wGE for the communities revealed that the whole brain wGE difference between conditions found in HDVs was entirely driven by the wGE of community 1. During Approach Alcohol, community 1’s wGE (0.321) was significantly higher (t(30) = 3.01, p = 0.005, Cohen’s d = 0.54) than during Avoid Alcohol wGE (0.303). Neither of the other two communities in HDVs showed a significant difference in wGE between conditions nor did any of the communities in HCs (Table 2). Only community 1 showed differences in wGE between conditions, therefore, all following post-hoc analyses of subcommunity structure were constrained on community 1.

### 3.7 Second Level Hierarchy: Subcommunity structure of Community 1

Whereas the 3-communities structure on the top level showed only 8 – 14 % differences in ROI community affiliation across groups and conditions, the subcommunity structure of community 1 was distinctly different between the two groups and in the case of the HDVs also between conditions. The result of the community detection procedure for community 1 revealed a 3-subcommunity structure for both conditions in HCs. At second hierarchy level, the subcommunities mirrored well established intrinsic connectivity networks; community 1 in HCs could be subdivided into (1) a *fronto-parietal control network* (FPN) with bilateral middle frontal gyrus, superior frontal gyrus and parietal regions, and (2) into two DMN subsystems (Andrews-Hanna et al., 2010) a) the *midline-core default mode network* (DMN) with medial prefrontal cortex and posterior cingulate/precuneus as its most prominent parts and b) the *medial-temporal DMN* (Andrews-Hanna et al. 2010) consisting of ventral medial PFC regions together with orbitofrontal cortex, temporal pole regions and subcortical brain regions (nucleus caudatus, thalamus). The most obvious difference in subcommunity structure between HDVs and HCs was that the midline-core DMN was not <u>a single unit</u> as in HCs but divided into a separate anterior midline-core DMN subcommunity and a posterior midline-core DMN subcommunity in both conditions. Additionally, community 1 in HDVs had a 4-subcommunity structure. Apart from anterior and posterior DMN subcommunities, community 1 also had a FPN and a medial-temporal DMN subcommunity during Approach Alcohol. During Avoid Alcohol however, community 1 was structured into a 3-subcommunity structure with FPN subcommunity, posterior DMN subcommunity, and a new anterior DMN plus subcommunity that resulted from a merging of the anterior DMN with the medial-temporal DMN subcommunity into a single unit (see Figure 3).

**Figure 3.**
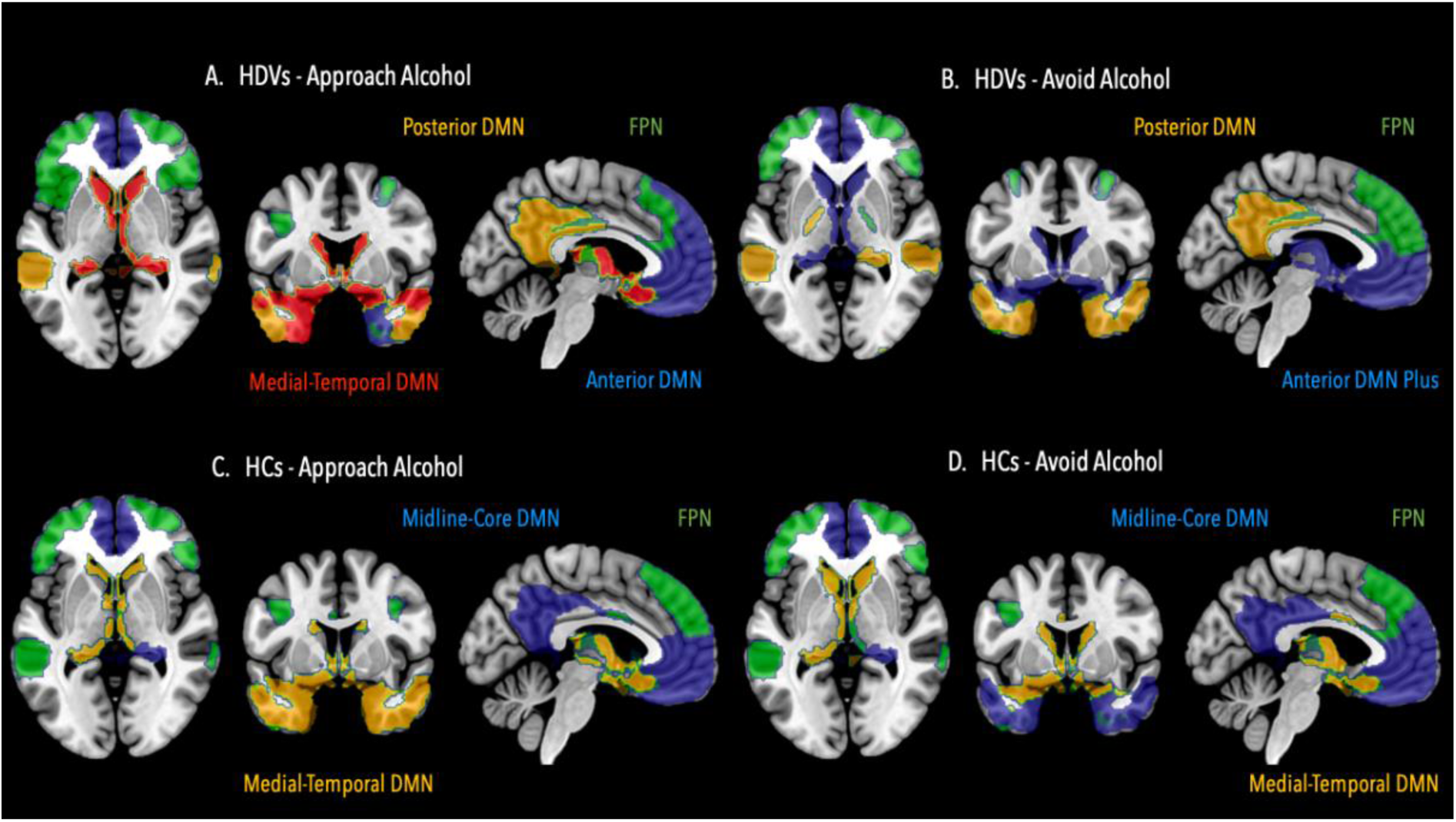
Second hierarchy level: Illustration of community 1’s subcommunity structure. Community 1 can be further partitioned into four subcommunities in heavy drinking veterans (HDV) for the Approach Alcohol condition (A); and into three subcommunities for the Avoid Alcohol condition (B). In healthy controls (HC), community 1 can be further partitioned into 3 subcommunities during the Approach Alcohol condition (C) as well as during the Avoid Alcohol condition (D). DMN = default mode network, FPN = fronto-parietal control network.

### 3.8 Second Level Hierarchy: Information integration between the subcommunities of community 1

In HCs, the two DMN subsystems (midline-core DMN and medial-temporal DMN) were characterized by significant information integration with each other during Approach Alcohol (ρ(17) = .770, p < .001) and Avoid Alcohol (ρ(17) = .533, p = .020) (Figure 4). However, there was additional significant inter-subcommunity information integration between midline-core DMN and FPN (ρ(17) = .528, p = .022) for Avoid Alcohol only, indicating a more prominent role of the FPN for inter-subcommunity information exchange during that condition. In HDVs, all four subcommunities of community 1 showed significant information integration between each other during Approach Alcohol (anterior DMN x FPN ρ(29) = .749, p < .001; anterior DMN x posterior DMN ρ(29) = .797, p < .001; anterior DMN x medial-temporal DMN ρ(29) = .560, p < .001; FPN x posterior DMN ρ(29) = .666, p < .001; FPN x medial-temporal DMN ρ(29) = .445, p < .013; posterior DMN x medial-temporal DMN ρ(29) = .719, p < .001). Information integration between the 3 subcommunities of community 1 during Avoid Alcohol however was reduced with only the posterior DMN having significant information integration with FPN (ρ(29) = .727, p < .001) and anterior DMN plus (ρ(29) = .383, p = .034).

**Figure 4.**
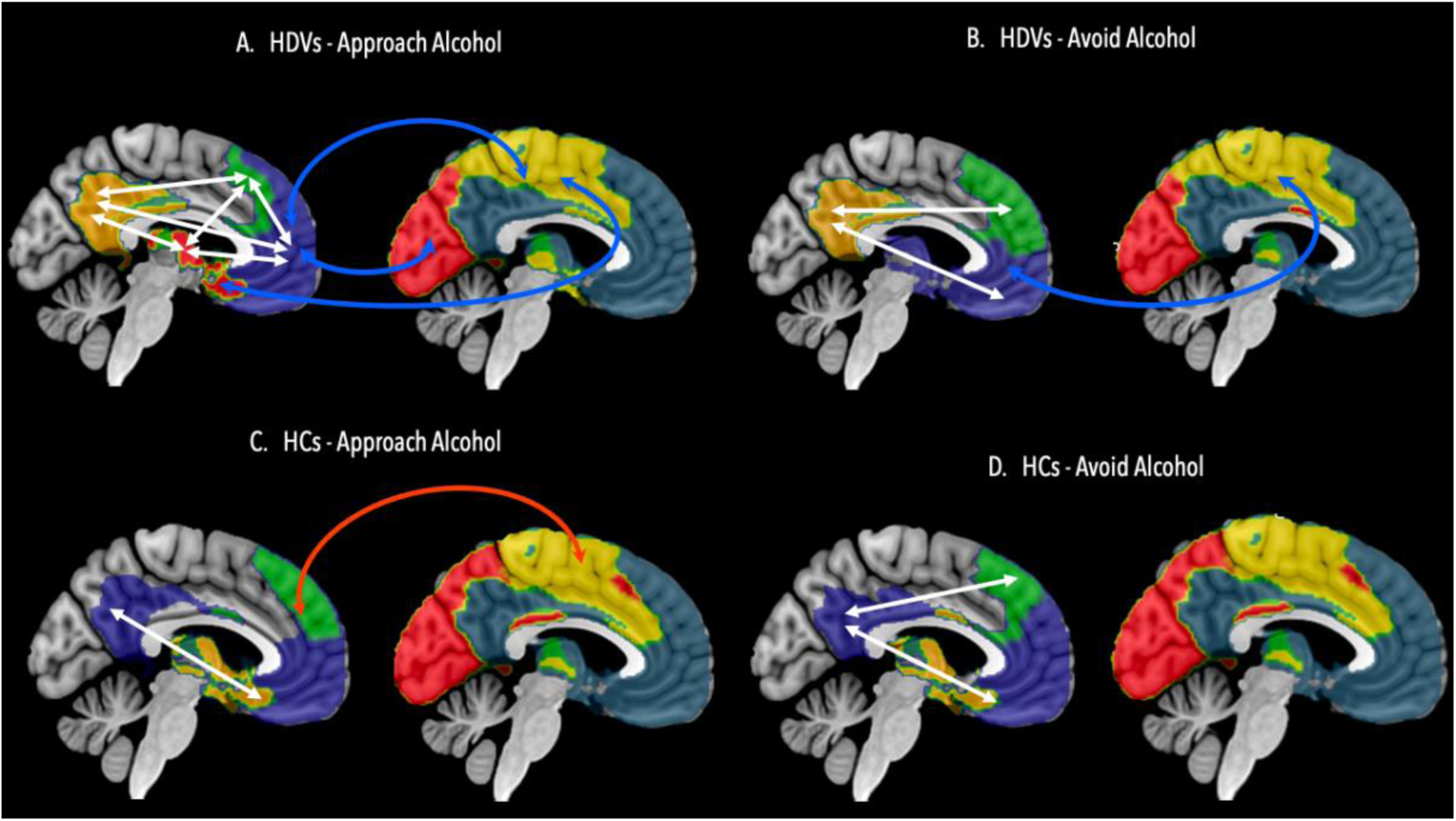
Illustration of significant information integration between community 1’s subcommunities and between the subcommunities and community 2 and 3. Significant information integration between the subcommunities of community 1 (blue) is highlighted by white arrows, significant information integration between community 1’s subcommunities and community 2 (yellow) and community 3 (red) is highlighted in red arrows whereas significant decoupling of information exchange between subcommunities and communities 2 and 3 is highlighted in in blue arrows in heavy drinking veterans (HDV) during Approach Alcohol condition (A), in HDVs during Avoid Alcohol condition (B), in healthy veterans (HC) during Approach Alcohol condition, in HCs during Avoid Alcohol condition (D).

### 3.9 Second Level Hierarchy: CombPC and wGE values on subcommunity-level

In HCs, the midline-core DMN subcommunity during Approach Alcohol had a significantly higher combPC value (midline-core DMN = 0.108) than all other subcommunities (FPN = 0.035, Wilcoxon W 179 p < .001, r = .88; medial-temporal DMN = 0.046, Wilcoxon W 190, p < .001, r = 1) as well as the highest wGE value (midline-core DMN = 0.433) of all other subcommunities (FPN = 0.397, n.s., medial-temporal DMN = 0.264, Wilcoxon W 190, p < .001, r = 1) thereby highlighting the midline-core DMN as the most influential subcommunity in HCs’ community 1 during Approach Alcohol (see Table 2). However, this was not the case for Avoid Alcohol during which the FPN had the highest (but not significantly) combPC value (FPN = 0.062) of the 3 subcommunities (midline-core DMN = 0.058, medial-temporal DMN = 0.032) and wGE value (FPN = 0.431, midline-core DMN = 0.041, medial-temporal DMN = 0.258) and thereby qualified as the most influential subcommunity in HCs during Avoid Alcohol. Compared to Approach Alcohol, FPN’s combPC (Wilcoxon W 40, p = .026, r = .58) and wGE (Wilcoxon W 34, p 0 .012, r = .64) values significantly increased during Avoid Alcohol whereas the combPC (Wilcoxon W 160, p = .007, r = .68) value of the midline-core DMN significantly decreased.

In HDVs, the anterior DMN subcommunity was the most influential subcommunity during Approach Alcohol because it had the highest combPC value (anterior DMN = 0.219) of all four subcommunities, and significantly higher than FPN (= 0.174, Wilcoxon W 398, p = .004, r = .58) and medial-temporal DMN (= 0.066, Wilcoxon W 496, p < .001, r = 1) but not than posterior DMN (= 0.206) as well as significantly higher wGE values (anterior DMN = 0.529) than the other three subcommunities (FPN = 0.428, Wilcoxon W 445, p < .001, r = .79; posterior DMN = 0.451, Wilcoxon W 394, p = .003, r = 59; medial-temporal DMN = 0.321, Wilcoxon W 495, p < .001, r = .99). During Avoid Alcohol however, anterior DMN plus switched the role of the most influential subcommunity with posterior DMN. The latter had a significantly higher combPC value (posterior DMN = 0.118) than the other two subcommunities (FPN = 0.082, Wilcoxon W 45, p < .001, r = .82, anterior DMN plus = 0.022, Wilcoxon W 0, p < .001, r = 1) and a significantly higher wGE value (posterior DMN = 0.376) than anterior DMN plus (= 0.276, Wilcoxon W 1, p < .001, r = .99). The wGE value of FPN was slightly higher than posterior DMN’s value but not significantly. Compared to Approach Alcohol, HDVs showed a significant decrease in wGE and average combPC values across all subcommunities during Avoid Alcohol: anterior DMN/anterior DMN plus (wGE Wilcoxon W 495, p < .001, r = .99; combPC Wilcoxon W 496, p < .001, r = 1), posterior DMN (wGE Wilcoxon W 462, p < .001, r = .86; combPC Wilcoxon W 442, p < .001, r = .78), and FPN (Wilcoxon W 459, p < .001, r = .85; combPC Wilcoxon W 418, p < .001, r = .68).

### 3.10 Behavioral relevance of inter-subcommunity information integration

To gain insight in how the degree of inter-subcommunity information integration was relevant for understanding self-reported craving for alcohol, Spearman’s rho between HDVs’ subcommunities combPC values with OCDS was computed (see Table 3). OCDS total in HDVs was significantly associated with the combPC values of the FPN subcommunity (ρ(29) = -.361, p = .046) in Approach Alcohol as well as with the combPC values of the FPN subcommunity (ρ(29) = -.38, p = .035) and the posterior DMN community (ρ(29) = -.376, p = .037). The obsessive ODCS sub-score was significantly associated with the combPC values of the anterior DMN subcommunity (ρ(29) = -.43, p = .016), the FPN subcommunity (ρ(29) = -.46, p = .009), the posterior DMN subcommunity (ρ(29) = -.41, p = .023) in Approach Alcohol and also with the combPC values of the FPN subcommunity (ρ(29) = -.36, p = .044) and the posterior DMN subcommunity (ρ(29) = -.40, p = .025) in Avoid Alcohol. Across both conditions, a higher degree of intercommunity information integration, in particular of the FPN subcommunity with the two midline-core DMN subcommunities, was associated with lower craving.

**Table 3.** Information integration between community 1’s subcommunities is significantly associated with OCDS craving scores in HDVs.

| Condition | OCDS Total | OCDS obsessive |
| --- | --- | --- |
| **Approach Alcohol** |  |  |
| Anterior DMN | – | $\rho = -.430$, $p = .016$ |
| Posterior DMN | – | $\rho = -.406$, $p = .023$ |
| FPN | $\rho = -.361$, $p = .046$ | $\rho = -.460$, $p = .009$ |
| **Avoid Alcohol** |  |  |
| Posterior DMN | $\rho = -.376$, $p = .036$ | $\rho = -.403$, $p = .025$ |
| FPN | $\rho = -.380$, $p = .035$ | $\rho = -.364$, $p = .044$ |

Craving as measured by the Obsessive Compulsive Drinking Scale (OCDS) is significantly associated with the degree information integration between the subcommunities in heavy drinking veterans during the conditions Approach Alcohol and Avoid Alcohol. DMN = default mode network and FPN = fronto-parietal control network

### 3.11 Information integration between subcommunities of community 1 with communities 2 and 3

Only one of the three subcommunities detected in HCs’ community 1 was found to have significant information integration with the other two communities. During Approach Alcohol but not during Avoid Alcohol, FPN showed significant information integration with community 2 (ρ(17) = .5, p = .031) and, on the level of a trend, i.e. the correlation coefficient would have reached significancy by applying a one-sided hypothesis, with community 3 (ρ(17) = .42, p = .078). HDVs differed from the HCs insofar that there was no significant information integration between the subcommunities of community 1 and the other two communities in HDVs but significant segregation: subcommunities anterior DMN and medial-temporal DMN showed significant segregation from community 2 and 3 during both conditions (for Approach Alcohol – anterior DMN x community 2 (ρ(29) = -.54, p = .002); anterior DMN x community 3 (ρ(29) = -.50, p =.005); medial-temporal DMN x community 2 (ρ(29) = - .41, p = .025; for Avoid Alcohol – anterior DMN plus x community 2 (ρ(29) = -.45, p = .011). In contrast to its frontal counterpart, subcommunity posterior DMN showed only minimal segregation from community 2 and even positive information integration with community 3, although not to a significant degree, during Approach Alcohol, and positive information integration, although again not significant, with both communities (see Figure 4).

## 4. Discussion

The study tested the hypothesis that AUD patients’ automatic action tendency to approach alcohol whenever presented with an alcohol-related stimulus has a neural signature in the functional whole-brain network architecture. We were able to confirm our hypothesis. HDVs had significantly higher wGE values for Approaching Alcohol than for Avoiding Alcohol which confirmed that their brain was more efficiently organized for approaching alcohol or “functionally set” to approach alcohol in the presence to alcohol-related external cues. In contrast, the HCs’ brains did not show such a processing advantage for either of the two action alternatives, their wGE values for the two conditions were almost identical. In a next step we wanted to explore which features of the HDVs functional network organization contributed to their processing advantage for approaching alcohol and how their functional network organization differed from the organization of the HCs. We were able to identify a set of frontal-parietally distributed brain regions grouped into a community encompassing the FPN and DMN during both conditions that was driving the HDVs’ higher functional efficiency for approaching alcohol. The same community also revealed characteristic differences in organization and information integration between HDVs and HCs. In the following sections, we will discuss these findings in more detail.

We start with the findings for HCs because describing the functional brain organization during the two alcohol conditions of the AAT in healthy brains allows us to set a point of reference to interpret the findings for the HDVs. HCs showed very similar community affiliations for both conditions on the top-level hierarchy as well as on the level of the subcommunities of community 1 indicating that their brains could switch between the two conditions without the need for extensive reconfiguration of the underlying network organization. Nevertheless, we observed significant differences in the degree of information integration globally as well as on the level of the communities and subcommunities during the two conditions. On the whole brain level, HCs showed significantly higher global whole brain PC values, and thereby a higher degree of global information integration, for Approach Alcohol. When interpreted within the framework of the global neural workspace theory (GNW; Dehaene et al., 1998; Finc et al., 2017, Kitzbichler et al., 2011, Vatansever et al., 2015), the finding of a higher degree of across community integration in HCs when approaching alcohol would indicate that this condition demanded relatively higher cognitive effort from the HCs than avoiding alcohol. According to the GNW theory, more demanding cognitive tasks require the engagement of a spatially more distributed and bigger set of brain regions than simple tasks. The three features of the HCs’ network organization during Approach Alcohol that contributed to this difference in global community integration fulfill the criteria of the global works space theory for more effortful processing. First, HCs showed significantly higher average combPC values for community 3 during Approach Alcohol. Second, although global information integration between community 2 and 3 was always significant, regardless of group or condition, between-community information exchange between these two communities as quantified by Spearman’s rho was significantly higher (p = .015) in HCs for Approach Alcohol than Avoid Alcohol. When combined and interpreted accordingly to the GNW theory, these two features would indicate a higher awareness of the visual stimulus characteristics in HCs during Approach Alcohol. Third and last, but probably in the context of the GNW theory most important, is the observation that the FPN subcommunity of community 1 showed significant information integration with community 2 suggesting a top-down influence of the higher-order FPN on that community which encompasses brain regions belonging to the ventral attention network, sensorimotor network and auditory-insula network.

Whereas the HCs’ brain configuration during Approach Alcohol revealed indicators that this condition was associated with relatively higher cognitive effort for them, the characteristics of the HCs’ functional network organization for Avoid Alcohol were, according to the GNW theory, more specific for tasks with relatively low cognitive demands. During Avoid Alcohol, HCs’ network organization showed a relative high degree of segregated processing typical for easy tasks or tasks that can be performed with some automaticity. Besides the already discussed finding of generally lower global whole brain PC and average community-level combPC values for the Avoid Alcohol versus Approach Alcohol, which is also indicative of more segregated processing when avoiding alcohol, the most obvious difference between the two conditions was the change of the FPN subcommunities engagement in within-community information integration. During Avoid Alcohol, the FPN subcommunity was no longer engaged in between-community information integration with community 2, but in significant information integration within its own community, particularly with the midline-core DMN subcommunity. As a result of the FPN’s switching from global between-communities to local within-community information integration when avoiding alcohol, the organization of the within-community information integration of community 1 was significantly affected. During Approach Alcohol, the midline-core DMN subcommunity was characterized by significantly higher wGE and combPC values compared to the other two subcommunities of community 1 which made the midline-core DMN the subcommunity with the strongest influence on the information integration or the “local leader” within community 1 during that condition. During Avoid Alcohol however, the FPN subcommunity overtook the role of a “local leader” of within-community information integration and was thereby having a top-down influence on two DMN subcommunities since it had significantly higher wGE and combPC values than the two other subcommunities.

In summary, HCs showed no such processing advantage in global wGE for either condition as we had observed for the HDVs. Nevertheless, Approach Alcohol seemed to be associated with relatively higher cognitive demand for them as indicated by the significant top-down information integration between the FPN subcommunity of community 1 and community 2. Switching between the two alcohol conditions of the AAT was characterized in HCs by different ways of FPN engagement with other communities and subcommunities. During Approach Alcohol, the FPN had the role of a “global player” because the subcommunity was engaged in significant information integration with community 2. In contrast, during Avoid Alcohol, the FPN showed the characteristics of a “local leader” because it was the most influential of the three subcommunities for within-community information integration. Speculatively one can wonder why the FPN in HCs was differently engaged in information integration during the two alcohol conditions. One possible explanation could be that HCs, light or non-drinking individuals, had to overcome their intrinsic tendency to avoid alcohol to the effect that approaching alcohol was more difficult for them and therefore required more cognitive control. Alternatively, HCs were less familiar with the alcohol related pictures and might have to exert more cognitive control to not get distracted and study the unfamiliar stimulus material but to keep their focus of attention on the direction of the tilt during Approach Alcohol.

On the top hierarchy level, the community structure of HDVs and HC was almost the same. Both groups showed a three-community structure for both conditions with only slightly different community affiliations. The only significant difference between the groups on this level was that the average combPC value for community 3 was significantly higher in HDVs when avoiding alcohol than in HCs. However, this significant difference between the two groups fits well with the general observation that the average combPC values of top-level communities showed the opposite information integration pattern in the two groups: whereas HDVs presented with overall higher average combPC values of the three communities during Avoid Alcohol, HCs average combPC values were generally higher during Approach Alcohol. When interpreted within the framework of the GNW theory, the higher degree of between-community information integration during Avoid Alcohol would indicate that avoiding alcohol was cognitively more effortful for the HDVs, which would also fit well with the fact that the HDVs’ whole brain functional network architecture showed a global processing advantage for approaching alcohol as measured by wGE.

Whereas the top hierarchy level of the community structure was quite similar between groups and conditions, this was no longer true for the subcommunity structure of community 1 which was different between the two groups so that comparisons on this level were no longer meaningful. The most obvious visual difference in the subcommunity structure of community 1 between HDVs and HCs was that the midline-core DMN in HDVs was not a single unit (as it had been in the HCs) but split into an anterior and posterior DMN. This subdivision of the midline-core DMN indicates that 1) the two core regions of the midline DMN had different roles in the between-community and between subcommunities information integration, and 2) their roles were different to such a degree that they were no longer forming a common subcommunity.

The primary aim of the study was to identify a neural signature associated with HDVs’ automatic action tendency to approach alcohol, so we now shift the discussion toward the findings for HDVs’ subcommunity organization of community 1 during Approach Alcohol. HDVs’ community 1 was organized into four separate subcommunities: anterior and posterior DMN, medial-temporal DMN and FPN. All four subcommunities were characterized by significant within-community information exchange between each other with the anterior DMN as the most influential subcommunity for integrating information between the four subcommunities or the “local leader” since it had significantly higher wGE and combPC values than the rest of the subcommunities. The inter-subcommunity information exchange was behaviorally relevant; a higher degree of inter-subcommunity information integration between three of the four subcommunities was significantly negatively associated with craving in HDVs (OCDS total score and “obsessive” subscale). The finding that craving, in particular obsessive thinking of alcohol, was significantly higher in HDVs with a lesser degree of inter-subcommunity information integration in community 1, is in line with the literature about rumination, which has repeatedly shown that higher DMN within-connectivity is associated with a higher degree of rumination (Li et al., 2018; Whitfield-Gabrieli & Ford 2012).

In contrast to HCs for whom we had observed significant top-down influence on information integration between FPN subcommunity and community 2 when approaching alcohol, HDVs showed significant levels of network segregation: anterior DMN as well as the medial-temporal DMN were significantly segregated from information integration with communities 2 and 3 during Approach Alcohol. Local processing by decoupling smaller groups of functional brain regions or subcommunities from the whole brain information integration is characteristic for tasks that can be performed automatically without much cognitive effort. The most prominent members of the anterior DMN subcommunity in HDVs were ventromedial prefrontal cortex and dorsomedial prefrontal cortex; the former is involved in computing expected value, reward outcome and experienced pleasure (Dohmatob et al., 2020, Doherty et al., 2015, Grabenhorst & Rolls, 2011, Vaidya & Badre, 2020), the latter is engaged during action consideration (Dohmatob et al., 2020; Kolling & O’Reilly, 2018). The observation that brain regions involved in reward evaluation and decision making were functionally segregated from brain regions involved in processing the visual stimuli and reacting to them could suggest that HDVs acted mainly “on decisional autopilot” when approaching alcohol.

When avoiding alcohol however, HDVs’ network organization showed signs of major re-organization since anterior DMN and medial-temporal DMN were merged into a single subcommunity (anterior DMN plus) during this condition whereas posterior DMN and FPN were still separate units. This re-organization was accompanied by changes in inter-subcommunity information integration: significant information integration existed only between posterior DMN and FPN and posterior DMN and the anterior DMN that was merged with the medial-temporal DMN into a single common subcommunity (= anterior DMN plus) when avoiding alcohol but no longer between anterior DMN plus and FPN. Additionally, during Avoid Alcohol, posterior DMN replaced the anterior DMN as the “local leader”, which had that role during Approach Alcohol, because the posterior DMN had significantly higher wGE and average combPC values than the other two subcommunities making it the most influential subcommunity for information integration. As for Approach Alcohol, organization of the inter-subcommunity information exchange was also behaviorally relevant during Avoid Alcohol: a higher degree of inter-subcommunity information integration was likewise associated with relatively reduced craving in posterior DMN and FPN but not in anterior DMN plus. In contrast to Approach Alcohol however, the capacity of the subcommunities for inter-subcommunity information integration as measured by their individual combPC values was significantly reduced during Avoid Alcohol. As a result, the internal subcommunity-organization of community 1 was less suited to control craving when avoiding alcohol than when approaching it.

According to the GNW theory, a higher level of global information integration is associated with higher cognitive effort and HDVs did indeed exert a higher degree of between-communities information integration during Avoid Alcohol: the average combPC values of all three communities were generally higher during Avoid Alcohol and the average combPC value of community 3 even significantly so. More important, community 3’s higher degree of information integration in HDVs was not caused by a higher degree of information exchange with community 2 as we had observed for the HCs during Approach Alcohol that was cognitively more demanding for them. In HDVs, Spearman’s rho quantifying the degree of information integration between two communities was even weaker for community 2 and 3 during Avoid Alcohol than Approach Alcohol, although not significantly weaker. The observed increase in between-community information integration for community 3 was with community 1; in particular with the posterior DMN subcommunity, which was the only subcommunity of community 1 with positive Spearman’s rho values (anterior DMN plus and FPN had both negative Spearman’s rho values and were therefore functionally separated from community 3) although none of the observed positive Spearman’s rho values for the posterior DMN subcommunity and community 3 was high enough to reach significance. This suggests that the posterior DMN subcommunity had an opposite role to its frontal midline-core DMN partner in HDVs when avoiding alcohol: instead of being segregated or decoupled it was integrated in the between-community information integration. The function of the posterior DMN with its key regions posterior cingulate, precuneus, and retrosplenial cortex within the brain network architecture is still quite an enigma (Cavanna & Trimble, 2006; Foster et al., 2023; Leech & Sharp, 2014; Leech & Smallwood 2019). One of the challenges in understanding the collective functional role of these brain regions interacting together as the posterior DMN is certainly that each of them separately can be further divided into smaller subunits with different connectivity profiles (Foster et al, 2023; Jiang et al., 2023; Leech et al., 2011; Leech et al, 2012; Willbrand et al., 2022) and these subunits dynamically change their network affiliation depending on the task at hand (Foster et al., 2023; Leech et al., 2011). The posterior cingulate cortex ROIs had the highest combPC values of all posterior DMN ROIs in HDVs indicating that this brain region primarily supported the observed between-community information integration exchange. Thus, we concentrate on the posterior cingulate cortex when interpreting the findings for the role of the posterior DMN in HDVs during Avoid Alcohol. Switching from a behavioral routine supported by a functional brain network organization that is optimized for implementing it with high automaticity to the behavioral opposite is effortful. Since it requires the brain not to just override old rules of how to act, but also to learn and implement new rules and reconfigure the brain network organization accordingly. The posterior cingulate cortex is a good candidate brain region for initializing and supporting this learning process as this region has been shown to be involved in change detection and altering or learning new behavior in response to changed circumstances where a current cognitive set or behavioral routine is no longer favorable and needs to be adapted accordingly (Dohmatob et al., 2020; Foster et al., 2023; Hayden et al., 2008; Pearson et al., 2011; Stawarczyk et al., 2021; Vatansever et al., 2017).

### 4.1 Limitations

Limitations include that the in-scanner version of the AAT is inherently different from the standard out of scanner behavioral AAT used to measure alcohol approach bias (e.g., limited response time window, variable intertrial intervals for fixed duration of overall task, and automatic advancement upon error). As such, we were unable to calculate simultaneous response times with enough accuracy to be used for correlation with fMRI brain outcomes. Additionally, the traditional AAT attempts to mimic naturally occurring approach and avoidance movements. However, the in-scanner AAT requires participants in supine body position with the joystick placed on the abdomen diminishing the natural arm extension and flexion movement. Ultimately, this positioning may have reduced the salience of the task and/or the associations that these behavioral reactions have with brain function. Another limitation is the somewhat small sample size, in particular of the HC group. However, fMRI data from healthy participants tends to have less variation and data quality is typically better. Despite the sample size, statistically significant effects were found. Additionally, most published studies investigating alcohol approach bias have been completed in non-treatment seeking or in residential treatment populations. Our study was focused on heavy drinking AUD patients engaged in outpatient treatment. Therefore, our findings may not be directly comparable to most of the published literature regarding alcohol approach bias among non-treatment seeking patients or those engaged in residential treatment. However, this is simultaneously a strength in that outpatient populations are not well represented in current research on alcohol approach bias, and these results extend work in this area.

### 4.2 Conclusion

HDVs’ functional brain organization was significantly more efficient organized when approaching alcohol compared to avoiding it. This functional processing advantage was characterized by how FPN and different DMN subsystems exchanged information with each other, by significant segregation of the anterior DMN, which is engaged in computing reward outcome and in assessing of the outcome of one’s own actions (Dohmatob et al., 2020), and from brain regions involved in implementing actions resulting from these decisional processes. Despite their functional brain organizations showing clear indicators of higher cognitive effort when HDVs were required to avoid alcohol, they were not completely able to override their brain’s “decisional autopilot” to approach alcohol, i.e., the anterior DMN continued to be characterized by functional segregation from the rest of the brain. However, there was also indication that the HDVs’ brain was trying to influence its internal model of how to respond when confronted with alcohol related context; the posterior cingulate cortex, which has been shown to be involved in global monitoring and information integration for influencing ongoing learning and decisions (Dohmatob et al., 2020, Foster et al., 2023) was found to have a significant degree of information integration with the FPN while also engaged (although not significantly) in information integration with the visual cortex. Results contribute to understanding the complex neural underpinnings of alcohol approach bias, emphasizing the involvement of FPN and DMN subsystems and lay the foundation for developing more potent targeted interventions to modify these neural patterns in heavy drinking Veterans engaged in outpatient treatment for AUD.

## Data Availability

All data produced in the present study are available upon reasonable request to the authors

## Acknowledgements

This work was supported by Department of VA Clinical Science Research & Development: IK2CX001510 (Pennington); National Institutes of Health: DA039903 (Pennington); and by San Francisco VA Healthcare System resources.

We extend our appreciation to all who volunteered for this research. We thank the Addiction and Recovery Treatment Service personnel at the San Francisco VA Health Care System for critical help with participant recruitment. We also wish to thank Brooke Lasher, Erica Walker, Alen Tersakyan (Add and Renée Dembo for managing and coordinating this clinical research project and for MR data acquisition.

## Conflict of Interest

The authors declare no conflict of interest.

## Author Contributions

VM writing (review and editing); AMM methodology, formal analysis, writing (original draft preparation, reviewing, and editing), visualization; DLP conceptualization, funding acquisition, supervision, project administration, writing (review and editing); CYFW (investigation)

## Notes

### Competing Interest Statement

The authors have declared no competing interest.

### Author Declarations

Ethics committee/IRB of the University of California, San Francisco and Ethics committee/IRB of the San Francisco Veterans Affairs Health Care System gave ethical approval for this work

